# An Agent Based Modeling of COVID-19: Validation, Analysis, and Recommendations

**DOI:** 10.1101/2020.07.05.20146977

**Authors:** Md. Salman Shamil, Farhanaz Farheen, Nabil Ibtehaz, Irtesam Mahmud Khan, M. Sohel Rahman

## Abstract

The Coronavirus disease 2019 (COVID-19) has resulted in an ongoing pandemic worldwide. Countries have adopted Non-pharmaceutical Interventions (NPI) to slow down the spread. This study proposes an Agent Based Model that simulates the spread of COVID-19 among the inhabitants of a city. The Agent Based Model can be accommodated for any location by integrating parameters specific to the city. The simulation gives the number of daily confirmed cases. Considering each person as an agent susceptible to COVID-19, the model causes infected individuals to transmit the disease via various actions performed every hour. The model is validated by comparing the simulation to the real data of Ford county, Kansas, USA. Different interventions including contact tracing are applied on a scaled down version of New York city, USA and the parameters that lead to a controlled epidemic are determined. Our experiments suggest that contact tracing via smartphones with more than 60% of the population owning a smartphone combined with a city-wide lock-down results in the effective reproduction number (*R*_*t*_) to fall below 1 within three weeks of intervention. In the case of 75% or more smartphone users, new infections are eliminated and the spread is contained within three months of intervention. Contact tracing accompanied with early lock-down can suppress the epidemic growth of COVID-19 completely with sufficient smartphone owners. In places where it is difficult to ensure a high percentage of smartphone ownership, tracing only emergency service providers during a lock-down can go a long way to contain the spread. No particular funding was available for this project.

## 1 Introduction

COVID-19 is a highly transmissible disease that was declared to be a global pandemic on March 11, 2020,^1^ by the World Health Organization (WHO). The disease is caused by a strain of Coronavirus, namely, Severe acute respiratory syndrome coronavirus 2 (SARS-CoV-2),^2^ and is highly infectious, resulting in a total of more than 8 million cases worldwide as of June 15, 2020.^3^ While originating in Wuhan, China in December, 2019,^4^ the disease has spread to more than 200 countries of the world by now.^3^ Countries have taken restrictive measures (i.e., lock-down) as the primary approach to contain the outbreak of the virus since effective clinical measures are yet to be found.^5, 6^ This disease has heavily affected the economies of most countries leading to the global coronavirus recession or the Great Shutdown.^7^

The SARS-CoV-2 virus spreads through close contacts, i.e., when a susceptible person inhales droplets coming from an infected individual through coughing or sneezing.^8^ It can also infect people if they touch their eyes, nose, or mouth after having physical contact with contaminated surfaces.^8^ This nature of infection has given rise to various preventive practices that are Non-pharmaceutical interventions (NPI), such as, wearing masks, personal protective equipment (PPE), washing hands frequently, staying home, and avoiding gatherings.^9^

Here, we present an Agent Based Model (ABM) in order to simulate the propagation of COVID-19 among the inhabitants of a city. Our model can be adapted for any realistic scenario by incorporating appropriate parameters specific to the city under consideration. We also examine the impacts of protective measures and city-wide lock-down on the infection spread and determine the suitable parameters that help to contain the spread. Our experiments further explore the conditions under which the so called Digital Herd Immunity,^10^ can be achieved by applying contact tracing approach via smartphones. Our results suggest that, with lock-down in effect, if more than 60% of the population in a city are traceable through smartphones, effective reproduction number (*R*_*t*_) falls below 1 within three weeks of intervention. Moreover, 75% or more of the population owning a smartphone results in *R*_*t*_ *<* 1 sooner and the new infections are eliminated completely within three months of intervention.

## 2 Methodology

### 2.1 The Data and Assumptions

We use two categories of data in our model as follows (see supplementary Sections 3-5 for details).

#### 1. Location-specific data

a. We use the demographics of the inhabitants in a particular city (i.e., education, employment, life expectancy, percentage of individuals having different professions, and the nature and timing of various tasks performed by the people) and the data related to the number of transports and the average family size.
b. We also use the data related to COVID-19 disease, its spread among the population and the response of authorities. These include the number of daily new infections in the city and the day of announcement of restrictive policies or awareness measures.

To conduct our simulations, we have collected the above data for Ford county, Kansas,^11–20^ and New York city,^21–27^ of United States of America.

#### 2. Physiological data

The probability of a person coughing and sneezing, touching contaminated objects, coming into physical contact of others, or washing hands are also important parameters of our model which would differ based on whether a person is at work, home, or is hospitalized.

As an abstraction, we ignore the changes in population of the city within the period of our simulation, i.e., we ignore births, deaths, and migrations.

### 2.2 The Agents

In our ABM, each person is an agent and each agent is susceptible to COVID-19. We consider five possible states for a person at any particular time as follows.

1. Not-Infected or healthy (H)
2. Infected, not contagious, asymptomatic (N)
3. Infected, contagious and asymptomatic (A)
4. Infected, contagious and symptomatic (S)
5. Dead or Recovered (D)

Figure 1a presents the time interval between different stages of infection,^28, 29^ and Figure 1b demonstrates the state transitions.

**Figure 1:**
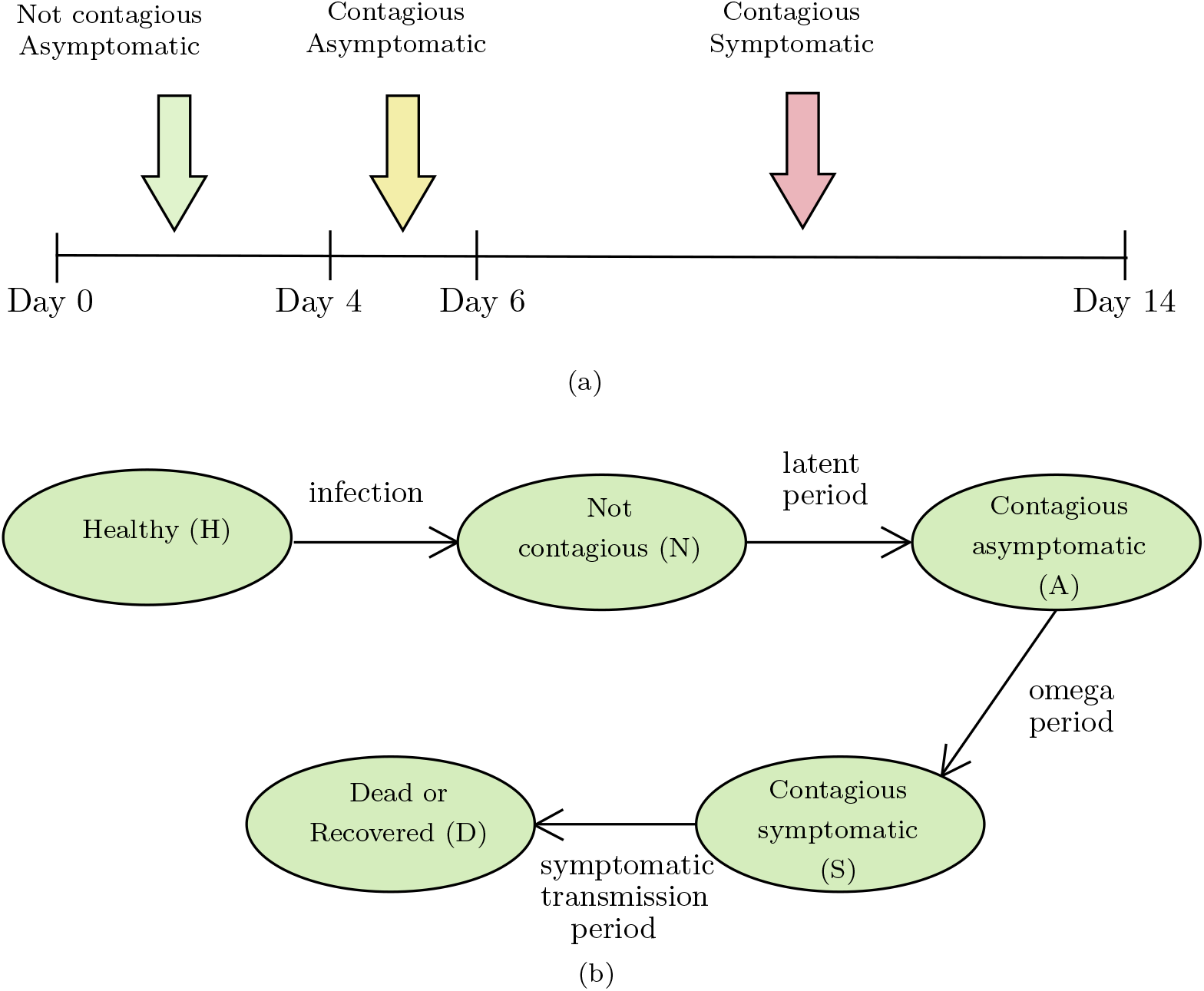
Stages of COVID-19 infection (1a) and state transitions in our model (1b).

Each agent is associated to a family and is assigned to one of four generic professions, namely, doctors and nurses (healthcare workers), students, service holders, and unemployed. The distribution and percentage of these professions are derived from real statistics. Based on his profession, an agent performs different actions throughout the day.

Each person has a *protection_level*. Once awareness is increased in the city, this value is improved with varying degrees. For doctors and healthcare professionals, a higher degree of protection is apportioned. The real-life significance of *protection_level* lies on the use of protective face shields, masks, maintaining hygiene etc.

### 2.3 Interaction between Agents and Transmission

Agents/persons in our model are associated to groups for interaction at every time unit (i.e., hour). Depending on whether the person’s task is to be at work, home, on a transport, in an event, or at a hospital during a particular hour, (s)he is assigned to a group. Every group, thus, involves a collection of people performing the following actions - sneezing and coughing, touching contaminated objects, coming into physical contact with each other, and washing hands. Only the action of washing hands has a positive impact and the impact is on himself whereas the rest take effect on others and the effect is negative. All these acts are given minimum and maximum probabilities of occurrence in an hour.

Within a group, agents can remain in different proximity with one another. Our model, at first, generates every possible pair of individuals in a group. Each pair then receives a numerical value (i.e., representing the proximity) chosen from one out of ten predefined ranges. Each range satisfies the relation 0 ≤ *B*_*low*_ < *B*_*high*_ ≤ 1 where *B*_*low*_ and *B*_*high*_ are lower and upper bounds of the range respectively.

The actions that lead to infection of another person will only matter when those are being executed by an infected person in the contagious stage of the disease. Equation 1 calculates the *infection* value of a susceptible person due to negative impacts of actions performed by an infected agent.

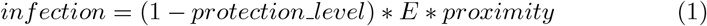

Here, 0 ≤ *O*_*min*_ ≤ *E* ≤ *O*_*max*_ ≤ 1; *O*_*min*_ and *O*_*max*_ represent lower and upper limits of the impacts of an infected person’s actions on others respectively.

On the other hand, equation 2 calculates the infection value due to positive effects of actions, i.e., washing hands.

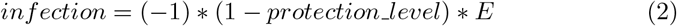

Here, *−*1 ≤ *S*_*min*_ ≤ *E* ≤ *S*_*max*_ ≤ 0; *S*_*min*_ and *S*_*max*_ are, respectively, the lower and upper limits of the impacts of an action on oneself.

This *infection* is then compared with an *action_infect_threshold*. Exceeding this threshold would cause the susceptible person’s *infection level* to be incremented (or decremented in case of actions with positive impact) by an amount equal to the *infection* value calculated above. By the end of the day, based on an *infection_threshold*, whether a person will be infected or not will be determined by this *infection level*. For more details about the model, see Supplementary Section 1.

### 2.4 Environment

#### 2.4.1 Awareness and Lock-down

This ABM involves a dynamic environment where factors change with time. A city may declare lock-down policies and awareness measures on the *X*^*th*^ and *Y* ^*th*^ days of community transmission. For the latter, a person’s *awareness level* is incremented from day *Y*. For a realistic interpretation, we allow some individuals to continue working or leaving their homes for different necessary tasks during lock-down.

#### 2.4.2 Contact Tracing and Quarantine

Traditional contact tracing through interviewing infected patients is not feasible.^10^ Leveraging smartphones through appropriate contact tracing apps,^30^ for tracking is a more reasonable option. Here we consider a scenario where all smartphone users would have contact tracing apps installed and will, thus, be brought under the umbrella of intervention. Our model traces (and subsequently quarantines for 14 days) someone who was in a group with an infected patient for a number of days prior to the onset of symptoms.

### 2.5 Model calibration, validation and simulations

To validate our model we run simulations (for a period of 60 days) of the spread of COVID-19 based on the data from Ford county as it has had a large number of tests and its data is relatively more available. Although an initial case of infection was identified on March 17, 2020 in Ford county, the infected individual was effectively isolated.^31^ We, thus, run the simulation from April 8 with 2 initial cases with lock-down imposed from day 1.^13^

Subsequently, we examine the effects of lock-down, contact tracing, and a combination thereof on a scaled down version of the New York City, United States of America and infer the factors that lead to digital herd immunity (See Supplementary Section 2). As for digital herd immunity, it has been found that there is always a critical fraction 0 ≤ *ϕ* _*c*_ *<* 1 of app ownership, such that take-up of contact-tracing apps by a fraction *ϕ > ϕ* _*c*_ of the population is sufficient to prevent epidemic spread.^10^ Finally, we apply these parameters to the simulation of Ford county. We further compute and assess the parameter called effective reproduction number (*R*_*t*_),^32^ following the method described in.^33^ If *R*_*t*_ *<* 1 can be sustained, it would signify that the number of new cases will gradually terminate among the population,^34^.^32^

### 2.6 Code and Availability

We have implemented our ABM and the experiments using Python3 programming language.^35^ The experiments have been conducted in the following machines - (i) a desktop computer having intel core i7-7700 processor (3.6 GHz, 8 MB cache) CPU, 16 GB RAM, and NVIDIA TITAN XP (12 GB, 1582 MHz) GPU; (ii) a Virtual Private Server (16 core CPU), 64 GB RAM and 200 GB Storage; (iii) a cloud computing platform Galileo from Hypernet (https://galileoapp.io/). All code and data can be found at the following link: https://github.com/s-shamil/agent-based-modeling-covid-19

## 3 Results

### 3.1 Model Validation using Ford County data

We ran the simulation for the full population (size = 33,619) of Ford county (Figure 2). The blue curve represents real data of daily confirmed cases. Our simulation results, represented by the red one, have a root mean square error (RMSE) of 50·2346 approximately (please see supplementary Table 5). The resulting curve is obtained by applying varying degrees of *protection_level* (defined in section 2.2), *action_infect_threshold*, and *infection_threshold* (defined in section 2.3) for different segments, i.e., ranges of days (Table 1 and Table 2).

**Figure 2:**
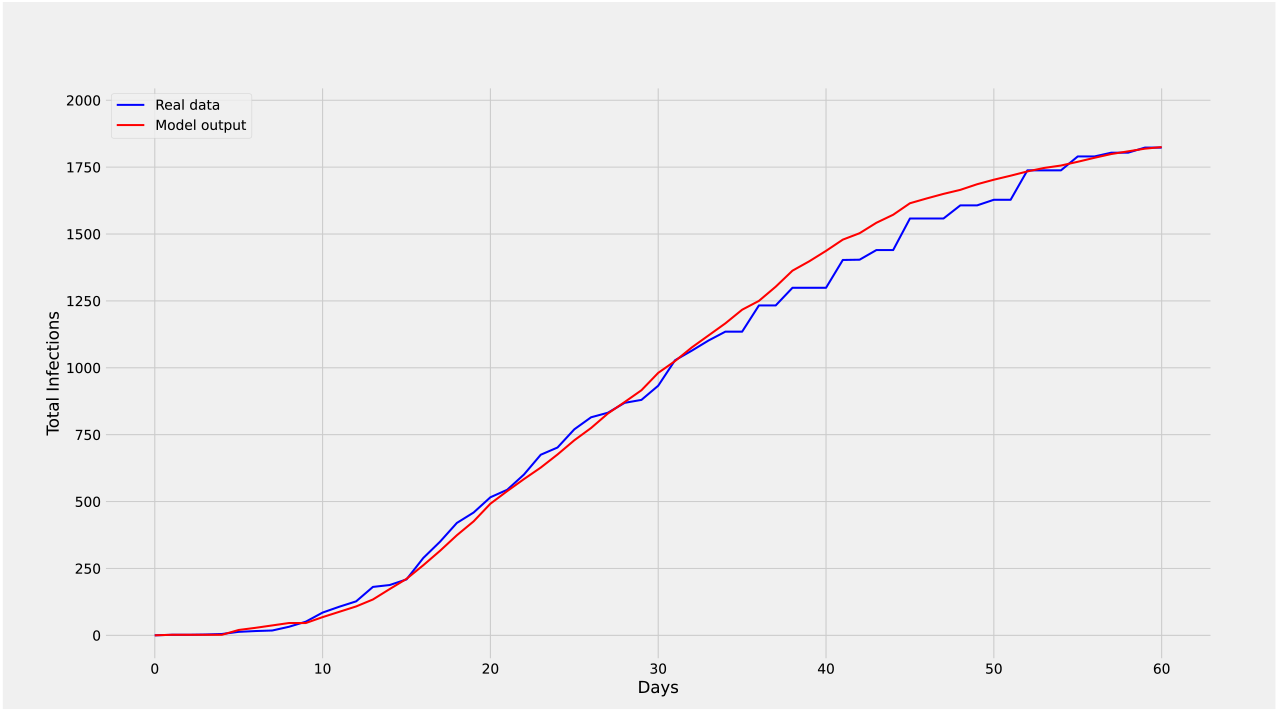
Validation of the model. The blue (red) curve represents the real (simulated) daily confirmed cases. The red curve nicely follows the blue curve with an RMSE of around 50·2346.

**Table 1:** *protection_level* on different days for inhabitants of Ford county. These values have been obtained through extensive experiments. Healthcare workers include all doctors, nurses, and people working in the health sector. Others include students, service holders, and unemployed people.

|  | Day 0-15 | Day 16-20 | Day 21-38 | Day 39-60 |
| --- | --- | --- | --- | --- |
| Healthcare workers | 0.2 | 0.22 | 0.8 | 0.95 |
| Others | 0.1 | 0.12 | 0.75 | 0.9 |

**Table 2:** *action_infect_threshold* and *infection_threshold* on different days for inhabitants of Ford county. These values have been obtained through extensive experiments.

|  | Day 0-8 | Day 9-20 | Day 21-38 | Day 39-45 | Day 46-60 |
| --- | --- | --- | --- | --- | --- |
| *action_infect_threshold* | 0.5 | 0.65 | 0.665 | 0.675 | 0.7 |
| *infection_threshold* | 0.5 | 0.65 | 0.65 | 0.65 | 0.65 |

### 3.2 Simulations on Ford County

#### 3.2.1 Effect of lesser interventions

We simulated to check how the outcome would change with a lesser degree of intervention. First, *protection_level* of the last segment (Day 39-60) was lowered by keeping it same as the previous segment (Day 21-38). This resulted in a 26·13% increase of daily incidence, whereas lifting movement restrictions from day 21 (as opposed to continuing the lock-down) results in an increase of 47 · 61% (Figure 3a). The variation of *R*_*t*_ for these changes are illustrated in Figure 3b.

**Figure 3:**
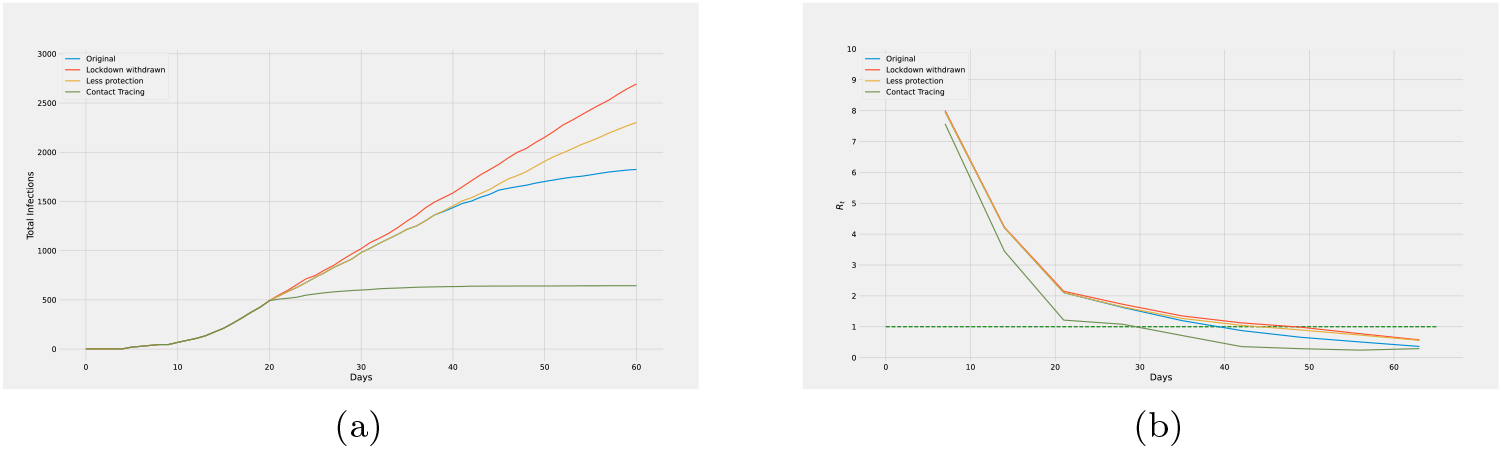
Effect of (i) removing lock-down from day 21 (red curve), (ii) allowing the *protection_level* of day 21-38 to continue until day 60 (yellow curve), and (iii) the simulation of daily confirmed cases (blue curve) are shown in 3a. Contact tracing from day 21 (green curve) with 75% smartphone users gives 3·51% less infections by the end of day 60 compared to the blue curve. Corresponding *R*_*t*_ curves are reported in 3b.

#### 3.2.2 Effect of the contract tracing

Figure 3a compares the impact of contact tracing implemented from day 21 (green curve) with the simulation of daily confirmed cases (blue curve). According to our simulation, the number of infections up to day 20 is 1 · 463% of the total population. In this simulation, we consider 75% of the people to be smartphone holders. However, not everyone in a group remains close enough (to the infected person) to allow smartphone applications to record their data. Considering 30% people in a group within reachable range and 90% group records being available, the simulation is continued till day 60 by tracing the contacts for the previous two days. Although infections continue to rise extensively in case of the blue curve, the green curve starts to flatten and the spread terminates with only 1·915% people being infected. The difference between the total infections for the two curves by the end of day 60 is about 3·51% of the entire population. As for the variation of *R*_*t*_ (Figure 3b), both the blue and green curves proceed to fall below 1 but the green one (i.e., involving contact tracing) does so sooner.

### 3.3 Simulation on New York City: A scaled down version

Simulation on New York city for a period of 120 days is done considering a population of 10000; parameters specific to New York City in the input are accommodated for the smaller population to ensure the appropriate scaling (see supplementary Table 11). As a basic validation of this scaled down version, the *R*_*t*_ values for the full population using an SIR model,^36^ and the same for the (scaled down) ABM model have been compared and the calculated RMSE was found to be 0·4626 (see section 6 of supplementary file for more details).

#### 3.3.1 Effect of Interventions

We have simulated four different scenarios with different intervention combinations, namely, no intervention (NI; blue), only contract tracing (CT; yellow), only lock-down (LD; red), and a combination of CT and LD (CT+LD; green). For all scenarios the parameters are same as NI till day 27 up to which only 5% people are infected.

We consider 75% of the agents to be traceable as smartphone owners.^37^ In a realistic scenario, lock-down policies and movement restrictions do not ensure that everyone will stay home. Thus, we consider that the lock-down will be effective on all students and on 50% of the service holders; among them, to simulate a practical lock-down, we keep 70% individuals strictly within their homes and allow the rest 30% to go out for different reasons. Evidently, CT+LD results in the least number of infections (Figure 4a). The relative trends of *R*_*t*_ also support this (Figure 4b) as *R*_*t*_ drops below 1 much faster for CT+LD.

**Figure 4:**
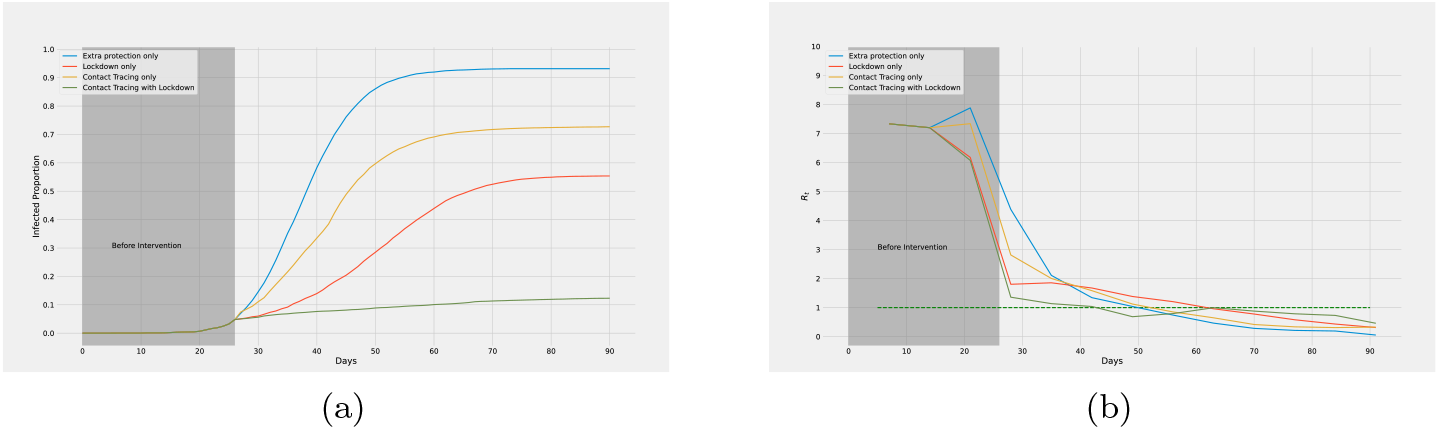
Figure 4a illustrates the effects of interventions imposed since day 27: introducing added protection (blue curve), contact tracing (CT) alone (yellow curve), lock-down (LD) alone (red curve), both contact tracing and lock-down (CT+LD) in combination (green curve). Corresponding *R*_*t*_ curves are illustrated in Figure 4b.

#### 3.3.2 Varying the percentage of smartphone users for contact tracing

Figure 5a illustrates a comparison considering different percentages of the population to be smartphone owners. In all these cases, we consider 30% people in a group associated with a person to be in close proximity and traceable only if they own smartphones. Moreover, among all the groups that a person stays in throughout the day, we take 90% of their records to be available. Figure 5b illustrates the corresponding variation in *R*_*t*_. As the percentage of smartphone owners increases, *R*_*t*_ falls below 1 more rapidly. Furthermore, Figure 5a demonstrates that the total percentage of population infected becomes much less with more ‘traceability’, i.e., with more fraction of smartphone users. Evidently, contact tracing with more than 60% smartphone users leads to *R*_*t*_ *<* 1 within three weeks of intervention. Moreover, new infections are eliminated completely among the population with more than 75% smartphone users as shown in Figure 5a within only three months of intervention.

**Figure 5:**
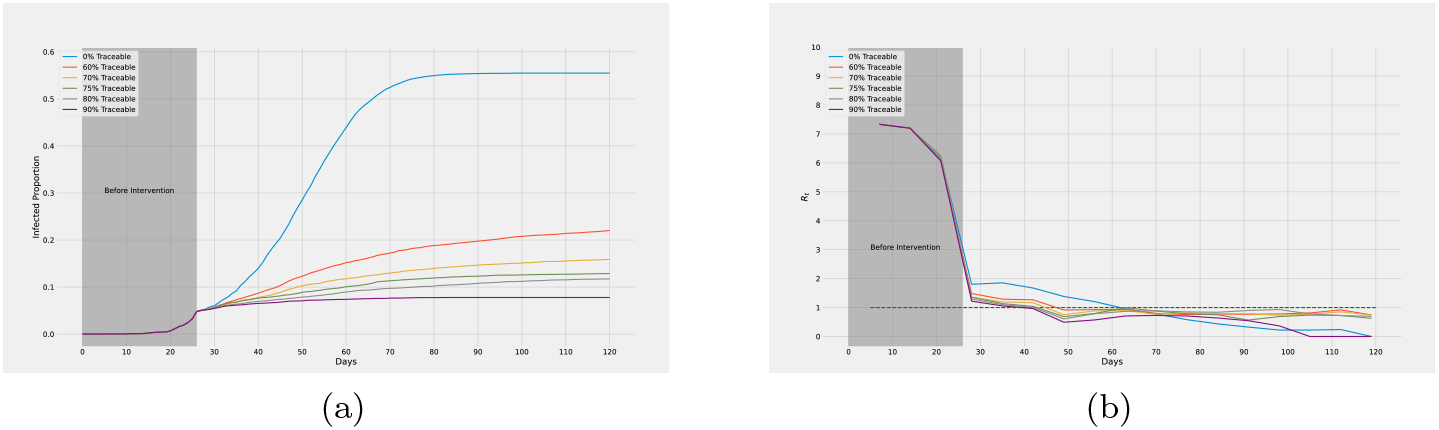
Introducing contact tracing with different percentage of the population being smartphone users from day 27. Figure 5a shows, with higher percentage of smartphone owners, less portion of the population are infected by the end of day 120. Variation of *R*_*t*_ is illustrated in Figure 5b.

#### 3.3.3 Combined impact of lock-down and contact tracing

Simulations were run to observe the impact of lock-down being initiated on different days while introducing contact tracing from day 27 in all cases (Figure 6a). By the end of 90 days, the curves having lock-down introduced on the 21st, 27th, and 41st days result in 3·36%, 12·87%, and 45·03% infections respectively. Clearly, an early announcement of lock-down regulation causes *R*_*t*_ to fall below 1 more quickly (Figure 6b).

**Figure 6:**
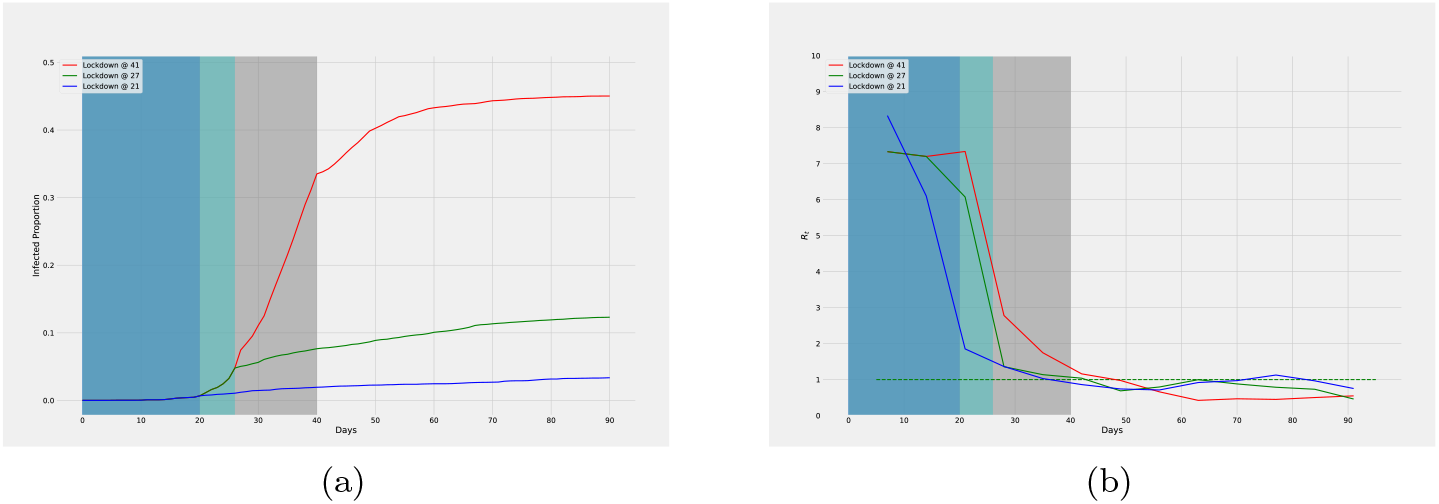
With contact tracing from day 27, Figure 6a shows effects of lock-down from day 21 (blue), 27 (green), and 41 (red). Variation of *R*_*t*_ is illustrated in Figure 6b.

Also, simulations were run to comprehend the effects of introducing contact tracing on different days with lock-down being declared on day 27 in all cases (Figure 7a). By the end of 90 days, the curves having contact tracing introduced on the 21st, 27th, and 41st days result in 8·72%, 12·87%, and 22·86% infections respectively. The corresponding graph for *R*_*t*_ is given in Figure 7b that shows similar results as in Figure 6b. However, compared to Figure 6b, the values of *R*_*t*_ take slightly longer to become less than 1.

**Figure 7:**
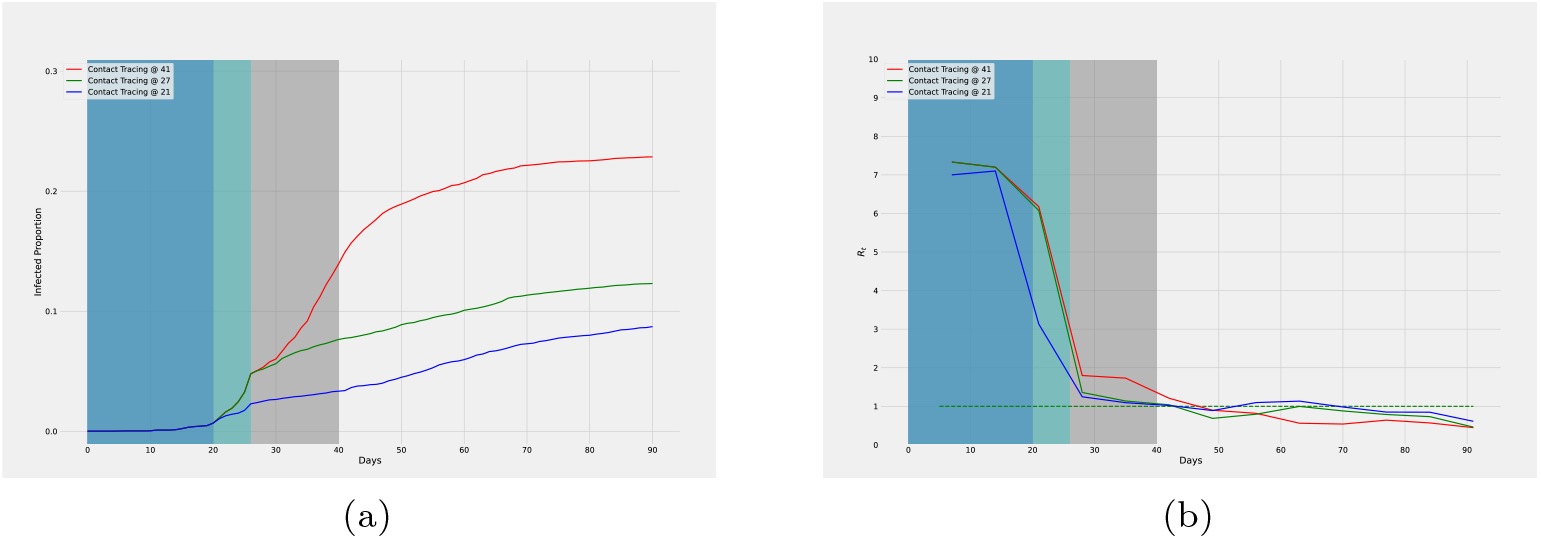
With lock-down from day 27, Figure 7a shows effects of contact tracing from day 21 (blue), 27 (green), and 41 (red). Variation of *R*_*t*_ is illustrated in Figure 7b.

#### 3.3.4 Some more variations

1. Tracing certain categories of people - Lock-down regulations only apply to all students and 50% of service holders. The rest of the people, i.e., doctors, nurses, healthcare workers, and a portion of service holders amount to about 40% of the population. Figure 8a illustrates a simulation where only the above-mentioned 40% population is traced (red curve) from day 27. Compared to the blue curve which illustrates tracing 75% of the entire population, tracing 40% causes 4· 58% less infections by the end of day 90. In both cases, contacts of the previous two days are traced for every infected person.
2. Effect of tracing for different number of days - In Figure 8a we show a comparison of our simulations conducted by allowing 75% smartphone users while tracing the contacts of the previous day only (shown in yellow). The blue curve (tracing the contacts for previous 2 days) results in 3· 45% less infections by the end of day 90.

**Figure 8:**
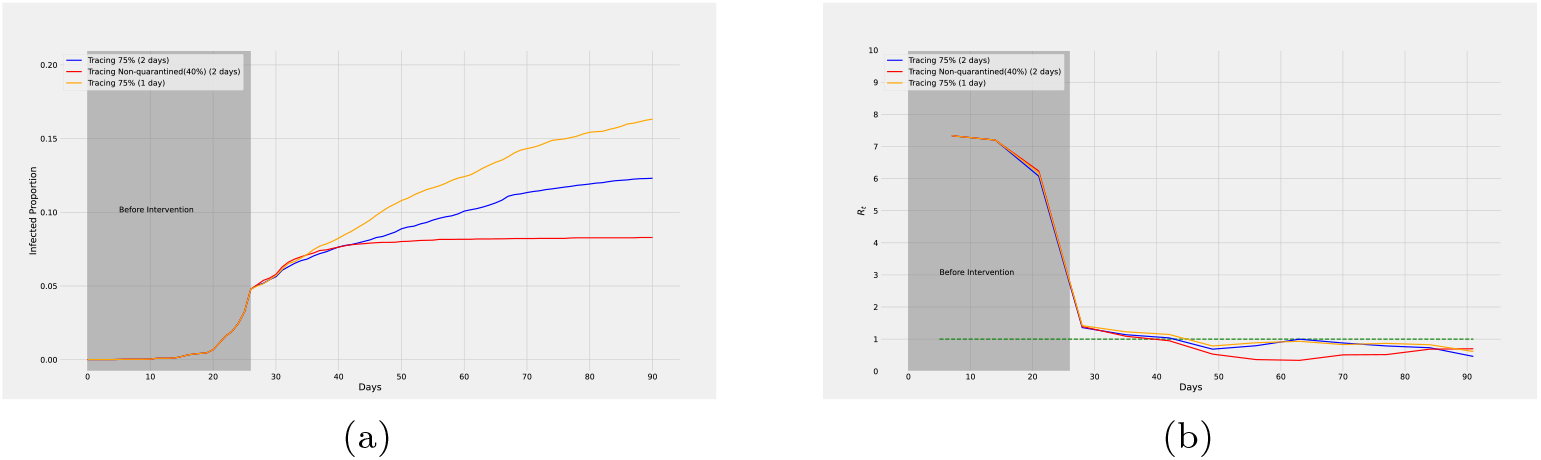
Figure 8a shows effects of tracing 75% of entire population (blue) as opposed to only the doctors, nurse, healthcare workers, and 50% service holders (red) from day 27. Both of these curves result from tracing the contacts of the previous two days. The yellow curve shows the effect of tracing 75% of the population for the previous day only. Their *R*_*t*_ curves are given in 8b.

## 4 Discussion

We found that only lock-down regulations can result in less number of people being infected in total compared to contact tracing approaches but it takes more time to reach *R*_*t*_ *<* 1. However, the success of combining lock-down and contact tracing surpasses all the other interventions significantly. This is evident from Figure 4a which shows that although the red curve (lock-down alone) causes 17 · 23% less infections than the yellow one (contract tracing alone), the green curve (both in combination) causes 42· 62% less infections than the red one.

Implementing contact tracing early will only work best if lock-down regulations follow shortly. With increasing delay in introducing these restrictions, the rate of infection rises more rapidly. This can be seen from Figure 6: although contact tracing has begun since day 27, the longer it takes to introduce lock-down following it, the greater is the increase in new infections everyday.

Lock-down regulations are the most effective when implemented early. Even if contact tracing measures are introduced late (red curve in Figure 7), they work well when supplemented with early lock-down as opposed to early contact tracing with delayed lock-down (red curve in Figure 6). This suggests that as long as lock-down is initiated early, delayed contact tracing will still work reasonably well. Ford county had issued movement restrictions from the beginning of community transmission;^13^ therefore, contact tracing works well in our simulation of the county (Figure 3).

Tracing a certain category of people may have a more profound effect on the overall disease spread. Effectively tracing individuals who go to their workplace, avail transport facilities, and attend small gatherings after lock-down regulations had already begun (i.e., doctors, nurses, healthcare workers, and about half of the service holders in our simulation reported in Figure 8) contains the spread with fewer total infections even if they represent a smaller fraction (only 40% in our simulation) of the susceptible population.

Owning smartphones (at varying percentage of the susceptible population) and thereby, ensuring (effective) contact tracing, ensures *R*_*t*_ *<* 1. Although not immediately, it does stop the spread of COVID-19 gradually. This will be quicker if lock-down is maintained. However, lock-down cannot be imposed for an indefinite period of time as it comes with economic repercussions. Therefore, it is important to know the fraction of smartphone owners in the susceptible population necessary to eliminate new infections very quickly. While having 60% or more smartphone owners in the susceptible population can slow down the spread, in order to completely contain it within a short period of time, this percentage should be maintained at 75% or more.

## 5 Conclusion

Our model can be adapted for simulating the spread of COVID-19 in any city by supplying suitable parameters as input. This can be used to observe what percentage of people in the city needs to remain traceable and for how many days in order to flatten the curve. This can also help gain insight on the future trend of the curve if current conditions persist for that city.

We have analyzed the effectiveness of digital herd immunity by exploring the impact of contact tracing and finding the parameters that lead to the termination of the epidemic within the city. Our results suggest that it is possible to reach digital herd immunity soon and with few new infections by implementing contact tracing as soon as possible if lock-down regulations had been implemented already. We have found from our experiments that, ensuring 75% smartphone owners in the population with at least 90% of the records being available in each person’s phone can completely contain the spread in a city within three months of intervention. Moreover, a more effective approach can be tracing all of the emergency service providers (who go to their workplaces) during the lock-down. Although they constitute only 40% of the population in our simulation, tracing only this category of people can flatten the curve of daily cases of COVID-19 within a short amount of time.

## Data Availability

All data and code are available at the following link:
https://github.com/s-shamil/agent-based-modeling-covid-19

## Acknowledgement

The authors deeply acknowledge the computing support provided by Brilliant Cloud Service (https://brilliant.com.bd/cloud) and Galileo from Hypernet (https://galileoapp.io/).

## Authors’ contributions

Shamil: Implemented the model, planned and designed the experiments, conducted the experiments, analyzed and interpreted the results, drafted the manuscript. Farheen: Implemented the model, planned and designed the experiments, conducted the experiments, analyzed and interpreted the results, drafted the manuscript. Ibtehaz: Implemented the model, planned and designed the experiments, analyzed and interpreted the results. Khan: Conducted some experiments. Rahman: Conceived the study, planned and designed the experiments, analyzed and interpreted the results, supervised the research work.

## Data and materials availability

All data and code are available at the following link: https://github.com/s-shamil/agent-based-modeling-covid-19

